# Context, Delivery Models, and Healthcare Outcomes of Community-Oriented Primary Care Services in Cuba: A Systematic Review of Narrative Evidence Protocol

**DOI:** 10.1101/2025.04.23.25326326

**Authors:** Yang Wang, Nuo Chen, Yi Wei, Dehua Yu, Jin Xu

## Abstract

**Objective:** This systematic review aims to summarize narratives related to the context, delivery models, and healthcare outcomes associated with Community-Oriented Primary Care (COPC) services provided by Cuban primary care practitioners to local community residents.

**Introduction:** “Community orientation” is a core feature of primary health care that enhances population health and a vital skill for general practitioners. COPC integrates primary health care and community medicine in a coordinated practice to improve population health. Cuba stands out globally, particularly among low- and middle-income countries, for its primary health care system and has achieved outstanding population health outcomes through this approach. However, systematic evidence summarizing the context, delivery models, and healthcare outcomes of COPC in Cuba remains lacking.

**Inclusion and Exclusion Criteria:** Included documents must: (1) focus on Cuban primary care practitioners and teams delivering community-oriented primary care services, along with local residents receiving and participating in these services; (2) explicitly address COPC services, including their context, delivery models, and healthcare outcomes; (3) be set in the Cuban community or primary care context; (4) consist of published or unpublished narratives within grey literature; (5) be in Spanish or English; and (6) be published on or after January 1, 1990. Excluded literature: (1) does not explicitly mention COPC services provided by Cuban practitioners, their context, or healthcare outcomes; (2) only addresses context beyond local health administration and community influence (e.g., U.S. economic embargo); (3) only involves events outside the Cuban primary care context (e.g., international medical missions); (4) is not a narrative (e.g., quantitative or qualitative studies).

**Methods:** We will search narrative literature published between January 1, 1990, and July 31, 2024, in Spanish or English across four academic databases: PubMed, Web of Science, Google Scholar, and Embase, and grey literature from the World Health Organization (WHO), the World Organization of Family Doctors (WONCA), including its Latin American branches, and the Cuban Ministry of Public Health websites. The review protocol is developed using JBI SUMARI. Qualified researchers will screen literature using Rayyan and JBI SUMARI, assess methodological quality, and extract data with JBI SUMARI tools. Data will be synthesized via the meta-aggregation method in JBI SUMARI.

**Systematic Review Registration Number:** This study is registered with PROSPERO, registration number is: CRD420251018485.

## Introduction

Primary care is characterized by a distinct set of functional features—first contact/accessibility, comprehensiveness, coordination, and continuity—crucial for improving health system performance, such as population health and controlling healthcare costs^1,2^. Among these, “community orientation” is key to integrating individual and population health interventions. World Organization of Family Doctors (WONCA) Europe defines^3^ it as “the ability of general practitioners to address the health needs of both individuals and the broader community based on available resources”. This ability depends on primary care physicians delivering primary care and public health services to local residents under resource-constrained conditions^4^, enabling them, as Muldoon et al^4^. noted, to “recognize and address social and environmental determinants of health through community knowledge and partnerships”.

One of the most well-known models that embody this feature is the model of Community-Oriented Primary Care (COPC)^5–7^. COPC operates under the principles of primary care, targeting clearly defined communities. It involves conducting community diagnoses, designing and implementing context-specific interventions, and monitoring their impact—all with active community participation^5,6,8^. According to the literature, health services closely tied to this model typically include screening, chronic disease management (e.g., hypertension and diabetes), and maternal and child health care^5^. This model has been implemented in various countries and regions, yielding notable positive impacts on health outcomes, such as blood pressure control in hypertensive patients, smoking cessation, and weight management^9–12^. However, its successful implementation is shaped by a complex interplay of external conditions, such as the availability of funding, supportive local government policies, the training and adaptability of general practitioners, and the degree of community organization and engagement^5,9–11^. There is an obvious knowledge gap about how COPC has worked in resource-constrained low- and middle-income countries (LMICs), where limited resources intersect with diverse and complex contextual factors.

Cuba stands out as one of the most prominent examples of LMICs that have prioritized primary care within its health system and considered one of the countries that exemplify COPC^12–14^. The country has also achieved population health outcomes that rival those of many developed countries, such as an infant mortality rate lower than that of the United States — often termed the “Cuban Miracle”^15–17^. These accomplishments are frequently attributed to its highly organized community participation and robust state-supported health policies. Yet, the reliability and generalizability of these achievements have been debated, with weak evidence quality and intricate health system dynamics complicating their interpretation^18–21^. This underscores the need to systematically evaluate the context, processes, and outcomes of Cuba’s COPC services. Insights from such an evaluation may be particularly valuable for countries like China, which are reforming their health systems to prioritize primary care, offering guidance on whether and how policymakers, administrators, and practitioners can adapt Cuba’s experience to shape effective policies, strategies, services, and clinical practices in resource-constrained settings.

Despite a clear need to systematically evaluate Cuba’s COPC services, such efforts remain largely absent from the existing literature. Preliminary searches on Google Scholar, PROSPERO, MEDLINE, the Cochrane Systematic Review Database, and the JBI Evidence Synthesis Database, using terms related to “COPC” (or “Primary Health Care”) and “Cuba”, revealed no completed or ongoing systematic reviews addressing this topic. This gap likely stems from the limited availability of robust quantitative and qualitative studies on Cuban primary care, compounded by the heterogeneous nature of the literature, which is dominated by narrative accounts. While these narratives offer rich descriptive detail about the community-oriented aspects of Cuban healthcare, their lack of standardized data, along with potentially diverged ideological perspectives, makes it challenging to synthesize coherent and actionable evidence. To address these challenges, this study adopts the Joanna Briggs Institute (JBI) methodology for systematic reviews of textual evidence, which is well-suited to structuring and synthesizing diverse narrative sources. This approach aims to clarify the context, delivery models, and health outcomes of Cuba’s COPC services, providing reliable insights for policy and practice in resource-constrained settings.

## Review questions

The questions of this review are:

1. What are the characteristics of the context of COPC in Cuba?
2. How are the COPC services organized and provided in Cuba?
3. What are the impacts of the COPC services in Cuba?

### Participants

Participants are Cuban primary care practitioners and teams delivering community-oriented primary care services, along with the local residents receiving and participating in these services.

### Phenomena of Interest

The phenomena of interest in this review are: (1) the context (e.g., policies, funding, or collaboration) closely related to COPC services, (2) the delivery models (e.g., service organization, community participation, or interventions) of these services, and (3) their healthcare outcomes (e.g., population health indicators, disease management, or patient satisfaction).

### Context

The context of this review is the primary care and community setting in Cuba.

### Types of publications

This review includes published narratives and unpublished narratives within grey literature, encompassing descriptive accounts of Cuban primary care experiences.

## Methods

This review will adhere to the Joanna Briggs Institute (JBI) methodological guidelines for systematic reviews of textual evidence^18^, focusing on synthesizing narratives related to Cuba’s community-oriented primary care services. The research protocol will be registered with PROSPERO (registration number: CRD420251018485). The review will follow this protocol, with any deviations transparently documented in the methodology section.

### Search Strategy

We have developed a three-stage systematic search strategy to identify published and unpublished narratives within grey literature relevant to the research questions. First, we will conduct dual-language (Spanish and English) searches based on three core concepts: “primary health care/general practice,” “community orientation/community-oriented primary care,” and “Cuba”—across four academic databases: PubMed, Web of Science, Google Scholar, and Embase (see Appendix I). Second, we will search grey literature on the websites of the World Health Organization (WHO), the Pan American Health Organization (PAHO), the World Organization of Family Doctors (WONCA), and the Cuban Ministry of Public Health. Third, during full-text screening, we will review the reference lists of included studies to uncover additional relevant literature.

Given that the primary language in Cuba is Spanish, we will limit the language of included literature to Spanish or English. To ensure both the timeliness of findings for contemporary relevance and the retrievability of records through modern databases, searches will cover literature published from January 1, 1990, to July 31, 2024. While this timeframe captures the modern evolution of Cuba’s COPC services, earlier foundational narratives may be considered if identified through reference tracking.

### Study selection

After completing the search, all identified literature will be uploaded to Rayyan for automatic and manual deduplication. An initial pilot screening will be conducted to refine the inclusion criteria and ensure consistency among reviewers. Subsequently, three reviewers (Yang Wang, Nuo Chen, and Yi Wei) will independently screen titles and abstracts in Rayyan based on predefined inclusion criteria. Full texts and bibliographic details of eligible studies will then undergo a second screening in Rayyan by the same three reviewers (Yang Wang, Nuo Chen, and Yi Wei). Yang Wang, who holds a medical degree from Havana Medical University in Cuba (six years of study with extensive engagement in local community health practices) and a Ph.D. in public health from Spain, is currently conducting primary care research in China. Nuo Chen and Yi Wei are students at Peking University, specializing in health policy research with experience in literature analysis and data extraction. Any disagreements during screening will be resolved through discussion among the three reviewers, with the results—including details of included and excluded studies—reported transparently in the final review using a PRISMA flow diagram^23^.

Included literature must meet the following criteria:

1. Participants are Cuban primary care practitioners and teams primary care services, along with local residents receiving and participating in these services;
2. The documents explicitly address the external conditions, delivery models, or healthcare outcomes of community-oriented primary care services;
3. The setting is the Cuban community or primary care context;
4. The sources are published narratives or unpublished narratives within grey literature;
5. The language is Spanish or English;
6. The publication date is on or after January 1, 1990.

Correspondingly, literature will be excluded based on the following criteria:

1. It does not explicitly mention community-oriented primary care services provided by Cuban practitioners, their context, or their potential healthcare outcomes;
2. It only addresses context beyond the influence of local health administration departments and community sectors (e.g., the U.S. economic embargo on Cuba);
3. It only involves events or issues outside the Cuban community or primary care context (e.g., Cuban medical missions abroad or the Latin American School of Medicine);
4. It is not a narrative, as defined by JBI (e.g., quantitative studies, qualitative studies, case studies, or literature reviews), but rather another form of descriptive account unrelated to the recounting of events in Cuban primary care.

Reasons for exclusion will be documented and reported in the final review.

Any disputes during the literature screening process will be resolved through discussion among the three reviewers (Yang Wang, Nuo Chen, and Yi Wei), reaching a consensus where possible. The results of the literature search, including details of included and excluded studies, will be reported transparently in the final review using a PRISMA flow diagram^23^.

### Assessment of methodological quality

Three independent reviewers (Yang Wang, Nuo Chen, and Yi Wei) will critically assess the methodological quality of the selected narratives based on the JBI critical appraisal guidelines for textual evidence, emphasizing authenticity as the primary criterion^24^. If necessary, we will contact the authors to clarify missing or additional data. Any disagreements will be resolved through discussion among the three reviewers, with input from Jin Xu as needed to facilitate consensus. The results of the critical appraisal will be presented in both narrative and tabular formats.

Following the JBI critical appraisal guidelines, we will exclude narratives from unreliable or inappropriate sources (Item 1), those where readers are unlikely to reach conclusions consistent with the authors’ (Item 4), and literature not recognized as narratives by the reviewers (Item 6). These exclusions will be determined after an initial quality assessment and group discussion involving Jin Xu, balancing comprehensiveness and rigor in the evidence obtained.

### Data extraction

Three reviewers (Yang Wang, Nuo Chen, and Yi Wei) will extract relevant data from studies meeting the methodological quality standards using the data extraction tools within JBI SUMARI. The extracted data will include specific populations, contextual settings, authors’ stated allegiance or position, conclusions regarding the three key phenomena (context, delivery models, and healthcare outcomes) with supporting text excerpts and page numbers, and the reviewers’ conclusions. Following the JBI methodological guidelines^22^, relevant content will be extracted in full paragraphs and assigned a credibility score (unequivocal, credible, or unsupported). If necessary, we will contact the authors to obtain missing or additional data, with Jin Xu providing input during discussions as required.

### Data synthesis

All textual data will be synthesized using the meta-aggregation method in JBI SUMARI^25^. We will adopt a three-tiered incremental synthesis approach: first summarizing key statements and main conclusions from the narratives, then categorizing them by similarity in meaning, and finally generating overarching statements representing distinct categories. These categories will undergo higher-level aggregation to produce overall findings supporting evidence-based practice. If meta-aggregation proves impractical (e.g., due to insufficient or highly heterogeneous narratives), results will be reported narratively. Only findings deemed unequivocal or credible will be included in the meta-aggregation.

### Assessing confidence in the findings

The JBI methodological guidelines for systematic reviews of textual evidence do not recommend using the GRADE or ConQual approaches for assessing certainty or confidence^20^. Given our study’s practical aim of providing background information to support the exploration and development of Community-Oriented Primary Care in contexts similar to Cuba’s, we will not evaluate the confidence of the synthesized data. Instead, we will present key findings descriptively in a narrative summary, enabling primary care researchers and practitioners to reference them according to their specific contexts.

## Data Availability

All data produced in the present study are available upon reasonable request to the authors

## Acknowledgements

None

## Author contributions

Conceptualization, Jin Xu and Yang Wang.; Methodology, Yang Wang.; Data curation, Yang Wang.; Formal analysis, Yang Wang, Yi Wei and Nuo Chen.; Funding acquisition, Jin Xu.; Project administration, Jin Xu. and Yang Wang.; Resources, Jin Xu and Yang Wang.; Supervision, Jin Xu.; Validation, Yang Wang, Jin Xu, Nuo Chen, Yi Wei and Dehua Yu; Writing—original draft, Yang Wang.; Writing—review and editing, Yang Wang, Yi Wei, Nuo Chen, Dehua Yu, and Jin Xu. All authors have read and agreed to the published version of the manuscript.

## Appendix

### Appendix I: Search strategy

#### PubMed

English: ((“family”[all fields] OR “family doctor”[all fields] OR “physician*”[all fields] OR “practice*”[tw] OR “primary care”[all fields] OR “Primary Health Care”[mh] OR “primary”[tw] OR “general pract*”[tiab] OR “COPC”[all fields] OR “policlínico”[all fields] OR “gp”[tiab] OR “gps”[tiab]) AND (“community”[all fields] OR “community-oriented”[all fields]) AND “cuba”[MeSH Terms])

Spanish: ((“familia”[all fields] OR “médico de familia”[all fields] OR “médico*”[all fields] OR “práctica*”[tw] OR “atención primaria”[all fields] OR “Atención Primaria de Salud”[all fields] OR “primaria”[tw] OR “medicina general”[tiab] OR “APOC”[all fields] OR “policlínico”[all fields] OR “médico general”[tiab] OR “médicos generales”[tiab]) AND (“comunidad”[all fields] OR “orientada a la comunidad”[all fields]) AND “Cuba”[MeSH Terms])

#### Web of Science

English: TS=(“primary care” OR “Primary Health Care” OR “primary health care” OR “general practice” OR “general practitioner” OR “family medicine” OR “family physician” OR “family doctor” OR “community-oriented primary care” OR “COPC” OR “policlínico”) AND TS=(“community” OR “community-oriented”) AND TS=(“Cuba”)

Spanish: TS=(“atención primaria” OR “Atención Primaria de Salud” OR “atención de salud primaria” OR “medicina general” OR “médico general” OR “medicina de familia” OR “médico de familia” OR “atención primaria orientada a la comunidad” OR “APOC” OR “policlínico”) AND TS=(“comunidad” OR “orientada a la comunidad”) AND TS=(“Cuba”)

#### Google Scholar (limited to the first 100 pages of results)

English: (“primary care” OR “Primary Health Care” OR “primary health care” OR “general practice” OR “general practitioner” OR “family medicine” OR “family physician” OR “family doctor” OR “community-oriented primary care” OR “COPC” OR “policlínico”) AND (“community” OR “community-oriented”) AND “Cuba”

Spanish: (“atención primaria” OR “Atención Primaria de Salud” OR “atención de salud primaria” OR “medicina general” OR “médico general” OR “medicina de familia” OR “médico de familia” OR “atención primaria orientada a la comunidad” OR “APOC” OR “policlínico”) AND (“comunidad” OR “orientada a la comunidad”) AND “Cuba”

#### Embase

English: (“family” OR “family doctor” OR “physician*” OR “practice*” OR “primary care” OR “primary health care”/exp OR “primary” OR “general pract*” OR “copc” OR “policlínico” OR “gp” OR “gps”) AND (“community” OR “community-oriented”) AND “cuba”/exp

Spanish: (“familia” OR “médico de familia” OR “médico*” OR “práctica*” OR “atención primaria” OR “atención primaria de salud”/exp OR “primaria” OR “medicina general” OR “apoc” OR “policlínico” OR “médico general” OR “médicos generales”) AND (“comunidad” OR “orientada a la comunidad”) AND “cuba”/exp

We will conduct searches on August 15, 2024, with publication dates restricted to January 1, 1990, through July 31, 2024, applied via database filters where available.

